# Gentamicin-Augmented Intrarenal Irrigation During Percutaneous Nephrolithotomy Reduces 30-Day Infectious Complications and Eliminates Pyelonephritis and Sepsis: A 1:2 Matched Cohort Study

**DOI:** 10.64898/2026.09.09.26362580

**Authors:** Pengbo Jiang, Kriselle Madamba, Jonathan Badin-Castro, Seyed Amiryaghoub Lavasani, Jocelyn Nguyen, Jaylen M. Lee, Bruce M. Gao, Sohrab N. Ali, Ryan S. Hsi, Roshan M. Patel, Jaime Landman, Ralph V. Clayman

## Abstract

**Purpose:** To evaluate whether gentamicin-augmented intrarenal irrigation during percutaneous nephrolithotomy (PCNL) was associated with fewer 30-day urinary infectious complications.

**Materials and Methods:** Single-center retrospective 1:2 matched cohort study of adults undergoing PCNL between January 2020 and June 2026. One hundred thirteen patients receiving gentamicin-augmented irrigation (80 mg per 3-L bag of 0.9% saline) were propensity-score matched to 226 saline controls on age, sex, body mass index, Charlson Comorbidity Index, ASA score, diabetes, and a pre-existing nephrostomy tube. The primary endpoint was a 30-day composite of symptomatic urinary tract infection with culture >1,000 CFU/mL, febrile UTI or pyelonephritis, or sepsis. A ≥100,000 CFU/mL composite was a prespecified sensitivity endpoint and asymptomatic bacteriuria a prespecified negative control.

**Results:** The primary composite infection outcome occurred in 3/113 (2.7%) versus 33/226 (14.6%) (risk difference −11.9%, 95% CI −17.4 to −5.6; NNT=8; p=0.001), and at ≥100,000 CFU in 1.8% versus 13.7% (p<0.001). Febrile UTI/pyelonephritis occurred in 0% versus 6.2% (p=0.006) and sepsis in 0% versus 4.0% (p=0.03), respectively. All events in the intervention arm were culture-defined UTI without fever or organ dysfunction. SIRS occurred in 31.9% versus 54.4% (p<0.001), while asymptomatic bacteriuria did not differ (2.7% versus 3.5%, p=0.758), respectively. Gentamicin remained protective after adjustment for demographic and stone characteristics (odds ratio 0.17, 95% CI 0.04–0.60; p=0.005). Acute kidney injury by postoperative day 1 occurred in 17.0% versus 17.6% (p=1.0).

**Conclusions:** Gentamicin-augmented intrarenal irrigation during PCNL was associated with fewer 30-day urinary infectious complications. Additionally, among treated patients meeting the primary outcome, there were no febrile or septic events, indicating a reduced severity of infectious complications when they did occur.

## Introduction

Percutaneous nephrolithotomy (PCNL) is the guideline-recommended treatment for large and complex renal calculi.^1^ Despite standard oral and parenteral antibiotic prophylaxis, postoperative urinary tract infection (UTI), systemic inflammatory response syndrome (SIRS), and urosepsis remain the most consequential complications of the procedure, with reported sepsis rates of 0.3–7.6%.^2,3^ Postoperative UTI and urosepsis after PCNL are estimated to add approximately $7,000 and $28,800 per event to the cost of care, respectively.^4,5^

Infectious risk after PCNL is multifactorial. A positive preoperative urine culture, diabetes mellitus, recurrent UTI, staghorn or high-burden stone disease, and indwelling urinary drainage are each independent predictors of post-PCNL SIRS and sepsis.^6,7^ The mechanism is attributed to bacteria residing within stone matrix and within the biofilm of indwelling stents and nephrostomy tubes, where penetration of systemically administered antibiotics is incomplete. Urothelial disruption and transient pyelo-venous and pyelo-lymphatic backflow during lithotripsy may then disseminate that bacterial burden into the circulation.^8–10^

Because PCNL requires continuous intrarenal irrigation, the irrigant itself is a potential route of drug delivery directly to the source of the bacteria. Previous work has shown that a povidone-iodine irrigation protocol sterilized the renal pelvis during PCNL, and antibiotic-augmented irrigation during retrograde intrarenal surgery reduced infectious complications in a prospective comparative study.^11,12^ We previously reported that intrarenal neomycin/polymyxin B irrigation was associated with markedly lower infectious morbidity in high-risk PCNL patients.^13^ That agent subsequently became commercially unavailable, and our institution adopted gentamicin at 80 mg per 3-L bag of 0.9% saline as its replacement. At that concentration (26.7 µg/mL) the irrigant exceeds the current CLSI susceptibility breakpoint for Enterobacterales by more than an order of magnitude, suggesting a bactericidal effect that may reduce infectious complications.^14^

We therefore evaluated whether gentamicin-augmented intrarenal irrigation during PCNL was associated with lower 30-day postoperative urinary infectious complications compared with standard saline irrigation.

## Materials and Methods

### Study Design and Ethical Oversight

This was a single-center retrospective matched cohort study of adults undergoing PCNL between January 2020 and June 2026 under an Institutional Review Board–approved protocol at the University of California, Irvine (IRB 2016-2746). The study period begins in January 2020 in order to keep the cohort within a similar surgical era and practice pattern; changes to the institutional picture archiving system and electronic medical record before that date also limited feasible chart review.

### Patient Selection, Treatment Assignment, and Matching

From our institutional percutaneous nephrolithotomy database we identified 499 PCNL procedures available for matching, of which 118 received gentamicin-augmented irrigation and 381 received saline alone. Gentamicin-augmented irrigation entered practice in July 2024 and was used at the discretion of the operating surgeon. Patients were grouped by receipt of gentamicin irrigation, and cases receiving neomycin/polymyxin B (GU) irrigant were excluded.

A propensity score for receipt of gentamicin irrigation was estimated by logistic regression from age, body mass index, sex, diabetes mellitus, American Society of Anesthesiologists (ASA) score, Charlson Comorbidity Index, and a pre-existing nephrostomy tube. Matching was 1:2 nearest-neighbor without replacement on the logit of the propensity score using a caliper of 0.2 standard deviations (width 0.18). Post-match balance was assessed using absolute standardized mean differences (SMD), with SMD ≥0.20 prespecified as meaningful imbalance.

### Perioperative Management

Procedures were performed by six endourologists at a single academic center. All positive preoperative urine cultures were treated with culture-directed antibiotics before surgery, and all patients received standard perioperative intravenous antibiotic prophylaxis. Intraoperative stone culture was obtained routinely. Upper-tract urine from the accessed kidney was not collected for culture in a standardized fashion before initiation of irrigation.

### Gentamicin Irrigation Protocol

Antibiotic-augmented irrigation was prepared by adding gentamicin 80 mg to each 3-L bag of 0.9% saline, yielding a concentration of 26.7 µg/mL. Control patients received 0.9% saline alone. Irrigation was administered using a Thermedx fluid management system (Thermedx, Solon, OH) at surgeon-selected pressures of 60–150 mmHg, delivered through the percutaneous access sheath with continuous egress to maintain a low-pressure collecting system. When retrograde access was employed, a ureteral access sheath was routinely used.

### Outcomes and Data Collection

Demographic, clinical, stone, operative, and postoperative data were abstracted from the medical record. The primary endpoint was a 30-day composite of (i) symptomatic UTI with a urine culture >1,000 CFU/mL, (ii) febrile UTI or pyelonephritis, or (iii) sepsis. A composite anchored at a urine culture threshold of ≥100,000 CFU/mL was prespecified as a sensitivity endpoint.

SIRS was defined by at least two of: temperature >38 °C or <36 °C, heart rate >90 beats/minute, respiratory rate >20 breaths/minute, or white blood cell count >12,000/µL or <4,000/µL. Sepsis was SIRS with a documented or clinically suspected urinary source. Sepsis-3 criteria were not applied because Sequential Organ Failure Assessment scoring was not recorded.^15,16^ Secondary outcomes were postoperative SIRS, 30-day emergency department visit for UTI, 30-day UTI-related readmission, and intensive care admission for sepsis.

Asymptomatic bacteriuria — a positive urine culture in the absence of urinary symptoms — was recorded separately and prespecified as a negative-control outcome.^17^

Preoperative stone burden was recorded as the largest stone dimension and as a stone volume, taken from computed tomographic segmentation where a segmented volume was recorded and calculated as an ellipsoid from the three maximal orthogonal dimensions otherwise. Postoperative non-contrast computed tomography was reviewed to assign standardized stone-free grades (Grade A, no residual; Grade B, ≤2 mm; Grade C, ≤4 mm) and to measure the largest residual fragment. Renal safety outcomes were serum creatinine change at postoperative day 0, day 1 and delayed follow-up, and KDIGO-defined acute kidney injury by postoperative day 1.^18^ In a subset of treated patients a postoperative serum gentamicin concentration was drawn at the treating team’s discretion as a safety check rather than as a pharmacokinetic protocol.

### Statistical Analysis

Continuous variables are summarized as mean ± standard deviation and categorical variables as frequency and percentage. Unadjusted comparisons used the Wilcoxon rank-sum, Fisher exact and chi-square tests as appropriate. Baseline balance was assessed by absolute standardized mean differences, with SMD ≥0.20 prespecified as meaningful imbalance. Risk differences are reported with Newcombe hybrid-score 95% confidence intervals, and number needed to treat only where that interval excluded zero.

Because the gentamicin arm contributed three primary events, all regression models used Firth penalized logistic regression, with penalized profile-likelihood confidence intervals and penalized likelihood-ratio tests.^19^ Model 1 used the covariate set from our prior irrigation study: treatment, age, diabetes, preoperative bacteriuria, a pre-existing nephrostomy tube, a pre-existing ureteral stent, infection stone composition and stone volume. Model 2 replaced infection stone composition with male sex, staghorn calculus, year of surgery and ultrasound-guided access. Prespecified sensitivity analyses restricted the cohort to surgery year 2024–2026, to a single high-volume surgeon, to the cohort with the two lowest-volume surgeons excluded, to ultrasound-guided cases, and to clinically defined subgroups; history of recurrent UTI was additionally entered into the adjusted model. Analyses were performed in R 4.3.3, with α = 0.05 and two-sided tests throughout.

### Economic Analysis

Lacking patient-level cost accounting, the economic component is a modeled estimate rather than a hospital cost analysis. Events prevented were calculated as the control-arm event rate applied to the 113 treated patients minus the events observed. Published U.S. attributable costs for sepsis hospitalization, infection-related readmission, emergency department visits and outpatient UTI treatment were applied and inflated to 2026 U.S. dollars using the Medical Care component of the Consumer Price Index.^4,5,20^ Gentamicin was costed at the Medicare Part B payment limit for 80 mg (HCPCS J1580), approximately $2.^21^ Unit costs were varied by ±20% in one-way sensitivity analyses.

## Results

### Cohort and baseline balance

Of the 118 gentamicin-eligible patients, 113 were matched and 5 were unmatched. The matched cohort comprised 339 PCNL procedures: 113 with gentamicin-augmented irrigation and 226 matched saline controls. Every propensity-score covariate balanced after matching, with a maximum absolute SMD of 0.07, as did variables not used in the match (Table 1). Age, sex, body mass index, Charlson Comorbidity Index, ASA score, diabetes, pre-existing urinary drainage, hypertension, chronic kidney disease stage, preoperative positive urine culture, staghorn calculus, largest stone dimension, stone volume, and number of stones all had SMD <0.20. Four variables exceeded that threshold. Any documented prior positive urine culture (44.2% versus 31.9%; SMD 0.258) and infection stone composition (62.8% versus 52.7%; SMD 0.205) were more common in the gentamicin arm; a pre-existing ureteral stent (12.4% versus 21.7%; SMD 0.239) and preoperative antibiotic administration (39.8% versus 50.0%; SMD 0.204) were less common.

**Table 1.** Baseline characteristics of matched percutaneous nephrolithotomy patients.

| Characteristic | Gentamicin irrigation (n=113) | Control (n=226) | SMD | p-value |
| --- | --- | --- | --- | --- |
| **Demographics** |  |  |  |  |
| Age, years (SD)<sup>†</sup> | 58.6 (14.0) | 58.4 (16.7) | 0.008 | 0.758 |
| Gender<sup>†</sup> |  |  | 0.036 | 0.844 |
| Female | 70 (62%) | 136 (60%) |  |  |
| Male | 43 (38%) | 90 (40%) |  |  |
| Body mass index, kg/m<sup>2</sup> (SD)<sup>†</sup> | 29.2 (7.3) | 29.2 (7.9) | 0.000 | 0.599 |
| **Clinical History** |  |  |  |  |
| Charlson Comorbidity Index (SD)<sup>†</sup> | 2.4 (1.8) | 2.4 (2.0) | 0.016 | 0.538 |
| ASA score (SD)<sup>†</sup> | 2.3 (0.6) | 2.3 (0.6) | 0.037 | 0.617 |
| Hypertension | 62 (55%) | 115 (51%) | 0.080 | 0.564 |
| Diabetes mellitus<sup>†</sup> | 32 (28%) | 57 (25%) | 0.070 | 0.601 |
| Chronic kidney disease* |  |  | 0.065 | 0.577 |
| Stage 1-2 | 88 (79%) | 163 (76%) |  |  |
| Stage 3 | 21 (19%) | 41 (19%) |  |  |
| Stage 4-5 | 3 (3%) | 11 (5%) |  |  |
| Recurrent UTIs | 50 (44%) | 72 (32%) | 0.258 | 0.031 |
| Any pre-existing urinary drainage tube | 30 (27%) | 73 (32%) | 0.125 | 0.317 |
| Ureteral stent | 14 (12%) | 49 (22%) | 0.239 | 0.039 |
| Nephrostomy tube<sup>†</sup> | 16 (14%) | 33 (15%) | 0.013 | 1.000 |
| Bladder catheter | 1 (1%) | 2 (1%) | 0.000 | 1.000 |
| Pre-operative positive urine culture | 30 (27%) | 57 (25%) | 0.030 | 0.793 |
| Pre-operative antibiotics | 45 (40%) | 113 (50%) | 0.204 | 0.084 |
| **Stone Characteristics** |  |  |  |  |
| Mean number of stones (SD) | 2.6 (2.0) | 2.3 (2.2) | 0.124 | 0.132 |
| Mean largest stone dimension, cm (SD) | 2.5 (1.3) | 2.4 (1.3) | 0.009 | 0.943 |
| Mean stone volume, cm<sup>3</sup> (SD) | 5.9 (9.5) | 6.6 (14.7) | 0.049 | 0.957 |
| Staghorn calculus | 47 (42%) | 87 (38%) | 0.063 | 0.638 |
| Infection stone composition | 71 (63%) | 119 (53%) | 0.205 | 0.082 |

SMD — standardized mean difference (absolute value). $p < 0.05$ indicates statistical significance.
<sup>†</sup>Variable entered into the propensity score used to match the gentamicin and saline groups. The full covariate set was age, body mass index, sex, diabetes, ASA score, Charlson Comorbidity Index and a pre-existing nephrostomy tube. Matching was 1:2 nearest-neighbor on the logit propensity score with a 0.2 SD caliper (width 0.18).
\*Chronic kidney disease stage derived from the CKD-EPI 2021 race-free equation using pre-operative creatinine, age and sex; categorized as stage $\geq 3$ versus 1-2 to calculate the SMD.

### Primary and secondary outcomes

The primary 30-day composite occurred in 3 of 113 gentamicin patients (2.7%) versus 33 of 226 controls (14.6%), a risk difference of −11.9 percentage points (95% CI −17.4 to −5.6), relative risk 0.18, number needed to treat 8 (p=0.001). At the prespecified ≥100,000 CFU/mL threshold the corresponding rates were 1.8% versus 13.7% (risk difference −11.9%, 95% CI −17.2 to −6.0; p<0.001). Symptomatic UTI with a positive culture occurred in 2.7% versus 12.4% (p=0.002) (Table 2).

**Table 2.** Post-operative urinary infectious complications of gentamicin irrigation and control groups.

| Outcome | Gentamicin<br>n/N (%) | Control n/N<br>(%) | Risk<br>difference | 95% CI | NNT | p-value |
| --- | --- | --- | --- | --- | --- | --- |
| **Post-operative UTI**<br>**composite (30 days)\*** | **3/113 (3%)** | **33/226**<br>**(15%)** | **-11.9%** | **-17.4% to -5.6%** | **8.4** | **0.001** |
| ≥100,000 CFU composite<br>(sensitivity) | 2/113 (2%) | 31/226 (14%) | -11.9% | -17.2% to -6.0% | 8.4 | <0.001 |
| Symptomatic UTI with positive<br>culture | 3/113 (3%) | 28/226 (12%) | -9.7% | -15.0% to -3.6% | 10.3 | 0.002 |
| Febrile UTI / pyelonephritis | 0/113 (0%) | 14/226 (6%) | -6.2% | -10.1% to -2.1% | 16.1 | 0.006 |
| Sepsis | 0/113 (0%) | 9/226 (4%) | -4.0% | -7.4% to -0.2% | 25.1 | 0.032 |
| Post-operative SIRS | 36/113 (32%) | 123/226<br>(54%) | -22.6% | -32.7% to -11.4% | 4.4 | <0.001 |
| Emergency department visit for<br>UTI (30 days) | 3/113 (3%) | 18/226 (8%) | -5.3% | -9.9% to 0.3% | — | 0.059 |
| Readmission for UTI (30 days) | 0/113 (0%) | 9/226 (4%) | -4.0% | -7.4% to -0.2% | 25.1 | 0.032 |
| ICU admission for sepsis | 0/113 (0%) | 3/226 (1%) | -1.3% | -3.8% to 2.1% | — | 0.554 |
| Asymptomatic bacteriuria<br>(secondary) | 3/113 (3%) | 8/226 (4%) | -0.9% | -4.6% to 4.3% | — | 0.758 |

NNT is shown only when the 95% CI for the risk difference excludes 0. $p < 0.05$ indicates statistical significance.
\*Composite of symptomatic urine culture >1,000 CFU/mL, febrile UTI or pyelonephritis, or sepsis within 30 days. Each patient is counted once; components may overlap.

Febrile UTI or pyelonephritis occurred in 0 of 113 gentamicin patients versus 14 of 226 controls (6.2%; p=0.006), and sepsis in 0 versus 9 (4.0%; p=0.032). Among the 33 control events, 9 were septic and a further 5 had febrile pyelonephritis without sepsis. All 3 events in the gentamicin arm were culture-defined UTI without fever or organ dysfunction. Postoperative SIRS occurred in 31.9% versus 54.4% (risk difference −22.6%, 95% CI −32.7 to −11.4; p<0.001). Emergency department visits for UTI (2.7% versus 8.0%; p=0.059), UTI-related readmission (0% versus 4.0%; p=0.032), and intensive care admission for sepsis (0% versus 1.3%; p=0.554).

Asymptomatic bacteriuria occurred in 2.7% of gentamicin patients versus 3.5% of controls (risk difference −0.9%, 95% CI −4.6 to 4.3; p=0.758).

### Adjusted analysis

In Model 1, gentamicin irrigation was independently associated with lower odds of the primary composite (adjusted OR 0.22, 95% CI 0.06–0.62; p=0.003). In Model 2, which additionally included male sex, staghorn calculus, year of surgery, and ultrasound-guided access, the association was unchanged (adjusted OR 0.17, 95% CI 0.04–0.60; p=0.005). Neither year of surgery (OR 1.16 per year, 95% CI 0.87–1.54; p=0.303) nor ultrasound-guided access (OR 0.71, 95% CI 0.22–2.17; p=0.546) was independently associated with infection, and neither was a pre-existing ureteral stent (OR 0.59, 95% CI 0.16–1.71; p=0.347) or stone volume (OR 1.01 per cm^3^, 95% CI 0.99–1.03; p=0.396). Male sex was protective (OR 0.07, 95% CI 0.01–0.30; p<0.001): 30 of the 33 control events occurred in women, with 1 event per 90 men (Table 3, Figure 1).

**Table 3.**
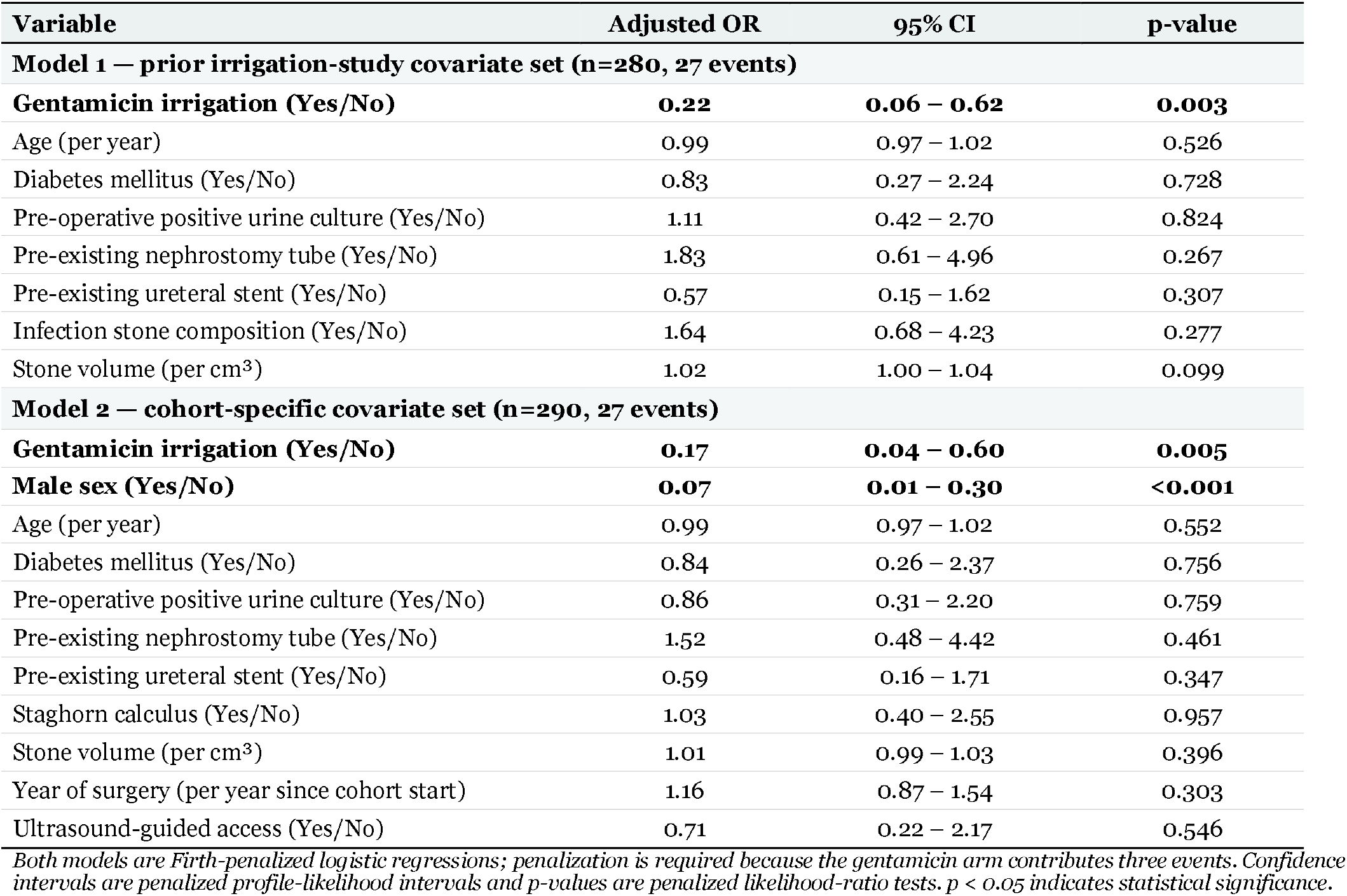
Multivariable logistic regression — predictors of 30-day post-operative UTI.

| Variable | Adjusted OR | 95% CI | p-value |
| --- | --- | --- | --- |
| **Model 1 — prior irrigation-study covariate set (n=280, 27 events)** |  |  |  |
| **Gentamicin irrigation (Yes/No)** | **0.22** | **0.06 – 0.62** | **0.003** |
| Age (per year) | 0.99 | 0.97 – 1.02 | 0.526 |
| Diabetes mellitus (Yes/No) | 0.83 | 0.27 – 2.24 | 0.728 |
| Pre-operative positive urine culture (Yes/No) | 1.11 | 0.42 – 2.70 | 0.824 |
| Pre-existing nephrostomy tube (Yes/No) | 1.83 | 0.61 – 4.96 | 0.267 |
| Pre-existing ureteral stent (Yes/No) | 0.57 | 0.15 – 1.62 | 0.307 |
| Infection stone composition (Yes/No) | 1.64 | 0.68 – 4.23 | 0.277 |
| Stone volume (per cm<sup>3</sup>) | 1.02 | 1.00 – 1.04 | 0.099 |
| **Model 2 — cohort-specific covariate set (n=290, 27 events)** |  |  |  |
| **Gentamicin irrigation (Yes/No)** | **0.17** | **0.04 – 0.60** | **0.005** |
| **Male sex (Yes/No)** | **0.07** | **0.01 – 0.30** | **<0.001** |
| Age (per year) | 0.99 | 0.97 – 1.02 | 0.552 |
| Diabetes mellitus (Yes/No) | 0.84 | 0.26 – 2.37 | 0.756 |
| Pre-operative positive urine culture (Yes/No) | 0.86 | 0.31 – 2.20 | 0.759 |
| Pre-existing nephrostomy tube (Yes/No) | 1.52 | 0.48 – 4.42 | 0.461 |
| Pre-existing ureteral stent (Yes/No) | 0.59 | 0.16 – 1.71 | 0.347 |
| Staghorn calculus (Yes/No) | 1.03 | 0.40 – 2.55 | 0.957 |
| Stone volume (per cm<sup>3</sup>) | 1.01 | 0.99 – 1.03 | 0.396 |
| Year of surgery (per year since cohort start) | 1.16 | 0.87 – 1.54 | 0.303 |
| Ultrasound-guided access (Yes/No) | 0.71 | 0.22 – 2.17 | 0.546 |

Both models are Firth-penalized logistic regressions; penalization is required because the gentamicin arm contributes three events. Confidence intervals are penalized profile-likelihood intervals and p-values are penalized likelihood-ratio tests. $p < 0.05$ indicates statistical significance.

**Figure 1.**
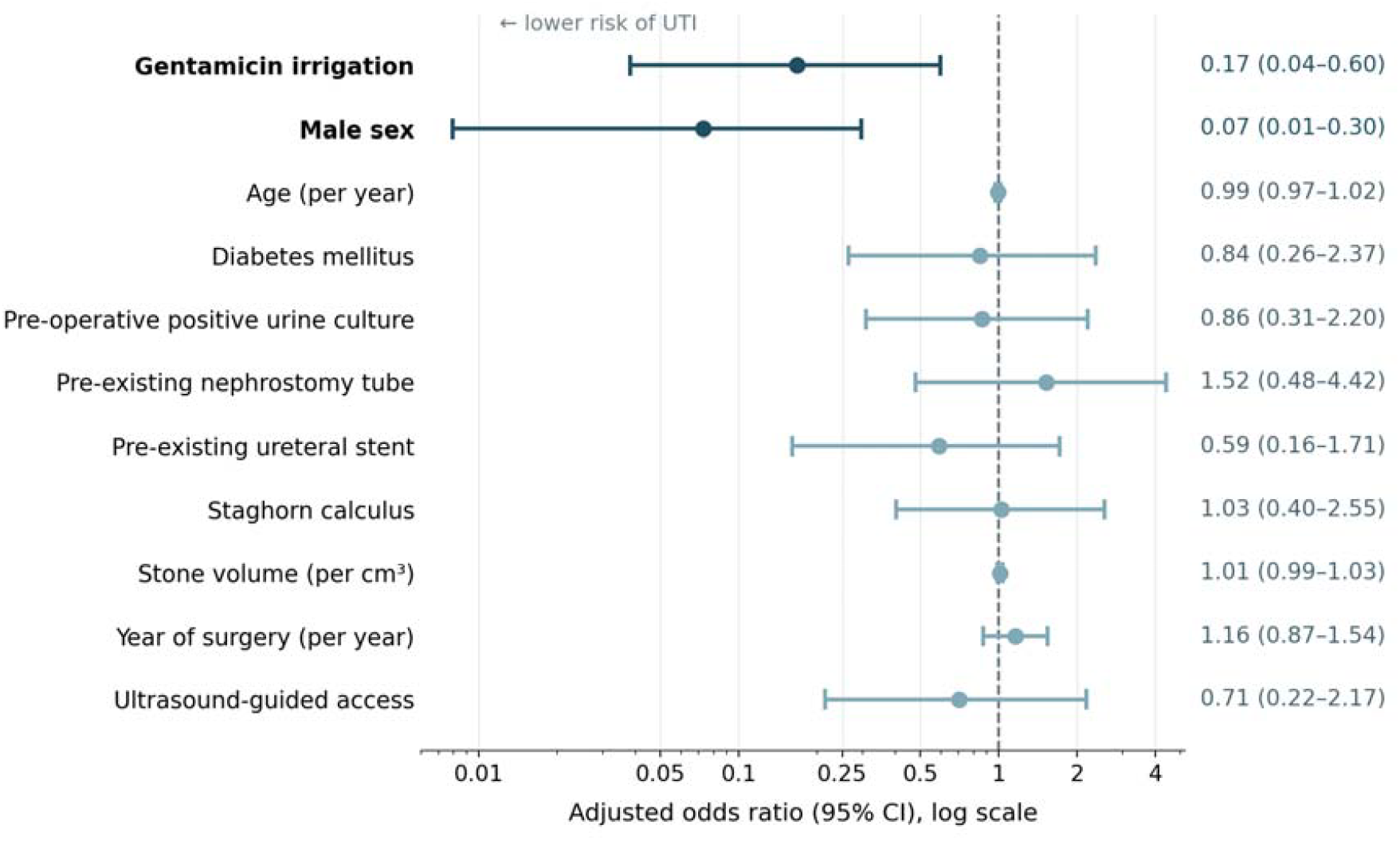
Forest plot of adjusted predictors of 30-day post-operative UTI Abbreviations: CI, confidence interval; OR, odds ratio. Adjusted odds ratios for 30-day post-operative UTI in the matched cohort (n = 290, 27 events), from Model 2. Covariates are gentamicin irrigation, male sex, age, diabetes, pre-operative positive urine culture, a pre-existing nephrostomy tube, a pre-existing ureteral stent, staghorn calculus, stone volume, year of surgery and ultrasound-guided access. Points represent odds ratios; horizontal bars show 95% penalized profile-likelihood confidence intervals; the dashed vertical line denotes an odds ratio of 1 (no effect).

### Sensitivity analyses

The association was stable across every prespecified sensitivity analysis. Restricting controls to the years 2024–2026 gave 2.7% versus 15.3% (relative risk 0.17; p=0.004), and to 2025–2026, 2.9% versus 22.2% (relative risk 0.13; p=0.001). Restriction to the single highest-volume surgeon gave 3.8% versus 15.1% (p=0.043), and exclusion of the two lowest-volume surgeons gave 2.8% versus 15.4% (p<0.001). Among ultrasound-guided cases only, rates were 2.9% versus 12.5% (p=0.098). The direction of effect was identical in all ten restrictions examined, including among patients with staghorn calculi (0% versus 21.8%; p<0.001) and among patients with a positive preoperative urine culture (3.3% versus 22.8%; p=0.029). Adding history of recurrent UTI to the adjusted model left the association intact (adjusted OR 0.18, 95% CI 0.04– 0.64; p=0.008).

### Perioperative and safety outcomes

Perioperative and safety findings are summarized in Table 4. Operative time was shorter in the gentamicin group (121.6 ± 56.0 versus 138.3 ± 64.8 minutes; p=0.025), as was length of stay (28.6 ± 18.7 versus 39.1 ± 37.3 hours; p=0.002). Postoperative leukocytosis was less frequent (36% versus 48%; p=0.038). Positive stone culture (48% versus 43%; p=0.487) did not differ.

**Table 4.** Operative characteristics and post-operative outcomes.

| Characteristic | Gentamicin irrigation (n=113) | Control (n=226) | p-value |
| --- | --- | --- | --- |
| **Operative Characteristics** |  |  |  |
| Laterality |  |  | 0.513 |
| Right | 57 (50%) | 104 (46%) |  |
| Left | 56 (50%) | 122 (54%) |  |
| Position |  |  | 0.521 |
| Prone | 98 (87%) | 201 (89%) |  |
| Supine | 15 (13%) | 23 (10%) |  |
| Access modality |  |  | <0.001 |
| Ultrasound | 69 (61%) | 40 (18%) |  |
| Fluoroscopy | 20 (18%) | 92 (41%) |  |
| Combined | 22 (19%) | 91 (41%) |  |
| Number of accesses (SD) | 1.01 (0.09) | 1.04 (0.22) | 0.143 |
| Stent placement (new) | 109 (96%) | 217 (96%) | 1.000 |
| Nephrostomy tube placement (new) | 12 (11%) | 28 (12%) | 0.723 |
| Bladder catheter placement | 104 (92%) | 197 (87%) | 0.205 |
| Mean case length, min (SD) | 121.6 (56.0) | 138.3 (64.8) | 0.025 |
| **Stone Characteristics** |  |  |  |
| Positive stone culture | 54 (48%) | 98 (43%) | 0.487 |
| Infection stone composition | 71 (63%) | 119 (53%) | 0.082 |
| **Post-Operative Characteristics** |  |  |  |
| Post-operative fever | 1 (1%) | 10 (4%) | 0.108 |
| Mean length of stay, hours (SD) | 28.6 (18.7) | 39.1 (37.3) | 0.002 |
| Blood transfusion | 0 (0%) | 1 (0.4%) | 1.000 |
| Post-operative leukocytosis ($>10.5 \times 10^3/\mu\text{L}$) | 41 (36%) | 109 (48%) | 0.038 |
| Mean change in WBC from preop ($10^3/\mu\text{L}$) (SD) | 2.1 (3.9) | 2.6 (4.7) | 0.087 |
| Mean change in serum creatinine |  |  |  |
| Preop □ POD 0 | +0.04 (0.19) | +0.02 (0.23) | 0.979 |
| Preop □ POD 1 | +0.08 (0.20) | +0.09 (0.31) | 0.947 |
| Preop □ delayed | +0.04 (0.27) | -0.01 (0.31) | 0.377 |
| # months postop, delayed creatinine | 2 (1-4); n=69 | 4 (2-6); n=113 | — |
| KDIGO AKI screen ($\Delta\text{Cr} \geq 0.3 \text{ mg/dL}$ by POD 1) | 19 (17.0%) | 37 (17.6%) | 1.000 |
| **Stone Clearance** |  |  |  |
| Stone-free grade (CT-assessed) |  |  | 0.084 |
| Grade A (absolute) | 49 (43%) | 76 (34%) |  |
| Grade B (0.1-2 mm) | 2 (2%) | 6 (3%) |  |
| Grade C (2.1-4 mm) | 5 (4%) | 23 (10%) |  |
| Not stone free ($>4 \text{ mm}$) | 39 (35%) | 99 (44%) | |
| Ungraded†† | 18 (16%) | 22 (10%) |  |
| Mean largest residual stone, mm (SD) | 4.9 (7.1) | 5.6 (6.7) | 0.119 |

$p < 0.05$ indicates statistical significance.
††Not graded because of unavailable or insufficient CT imaging, or a staged procedure with no interval imaging.

Mean change in serum creatinine from baseline did not differ at postoperative day 0 (+0.04 versus +0.02 mg/dL; p=0.979), day 1 (+0.08 versus +0.09; p=0.947), or delayed follow-up (+0.04 versus −0.01; p=0.377). KDIGO-defined acute kidney injury by postoperative day 1 occurred in 19 of 112 gentamicin patients (17.0%) versus 37 of 210 controls (17.6%; p=1.000). One transfusion occurred in a control patient. Postoperative serum gentamicin was measured in a subset of treated patients: concentrations were below the assay limit of quantitation (<0.3 µg/mL) in the majority of sampled operations, and no evaluable sample exceeded 0.6 µg/mL. No hypersensitivity reactions, neuromuscular events, or patient-reported hearing symptoms were documented.

Stone clearance did not differ between groups. Among the 299 patients with available postoperative CT, Grade A clearance was achieved in 49 of 95 gentamicin patients versus 76 of 204 controls, with similar distributions across grades (p=0.084 across the full grade distribution), and the mean largest residual fragment was 4.9 ± 7.1 versus 5.6 ± 6.7 mm (p=0.119).

### Economic analysis

Applying published U.S. unit costs to the events prevented across the 113 treated patients — 4.5 sepsis hospitalizations, 4.5 UTI-related readmissions, 6.0 emergency department visits, and 4.5 outpatient UTI episodes — yielded estimated gross savings of approximately $245,100, or roughly $2,200 per patient treated (Table 5). At roughly $2 per 3-L bag the irrigant adds under $25 per case even at high irrigation volumes, so net and gross savings are effectively identical. One-way variation of unit costs by ±20% did not change the direction of the modeled findings.

**Table 5.** Economic impact of gentamicin irrigation versus control (2026 USD)

| Resource / complication | Control rate | Expected events | Observed events | Events prevented | 2026 adjusted unit cost | Savings (USD) |
| --- | --- | --- | --- | --- | --- | --- |
| Sepsis (index hospitalization) | 4.0% | 4.5 | 0 | 4.5 | \$30,800 | \$138,600 |
| Readmission for UTI | 4.0% | 4.5 | 0 | 4.5 | \$19,600 | \$88,200 |
| Emergency department visit for UTI | 8.0% | 9.0 | 3 | 6.0 | \$2,800 | \$16,800 |
| Outpatient UTI treatment | 4.0% | 4.5 | 0 | 4.5 | \$300 | \$1,350 |
| **Total gross savings (113 treated patients)** | | | | | | **\$245,100** |
| **Approximate gross savings per patient treated** | | | | | | **\$2,200** |

*Because the arms are matched 1:2, events prevented are calculated as the control-arm event rate applied to the 113 treated patients minus the events actually observed, rather than as a raw count difference. Unit costs derive from nationally representative datasets and peer-reviewed studies, are inflated to 2026 U.S. dollars using the Medical Care component of the Consumer Price Index, and are rounded to the nearest hundred.*

## Discussion

In this matched cohort study, gentamicin-augmented intrarenal irrigation was associated with a substantial reduction in 30-day urinary infectious complications among patients undergoing PCNL. The effect was concentrated at the severe end of the spectrum. There were no febrile urinary infections and no septic events among 113 treated patients, compared to the rates of 6.2% and 4.0% in matched controls. The association persisted after adjustment and across the prespecified sensitivity analyses.

Three features of the comparison deserve comment. First, four baseline variables remained imbalanced after matching. Prior positive urine culture and infection stone composition were more common in the treated arm and preoperative antibiotics less common, all of which make the estimate conservative; the fourth, a pre-existing ureteral stent, was also less common in the treated arm, but it was entered directly in both adjusted models and was not independently associated with infection. Second, the treated and control arms did not fully overlap in the year of surgery. However, in adjusted analyses neither year of surgery nor method of percutaneous access was independently associated with infection, and restricting controls to 2024–2026 produced, if anything, a larger effect. Third, rate of asymptomatic bacteriuria was similar between groups (2.7% versus 3.5%), and positive stone cultures were no less frequent (48% versus 43%). This matters because it separates two explanations for the result: if the irrigant were simply sterilizing cultures rather than preventing infection, both should have fallen alongside the clinical endpoints — the stone in particular, since it is bathed in irrigant throughout the case. Neither did, which favors prevention of clinical infection over suppression of culture growth.

Gentamicin has been used as a urologic irrigant in other settings. Chronic gentamicin bladder irrigation has an established safety record in complex urologic patients, and intraoperative gentamicin bladder irrigation at the time of kidney transplantation was associated with fewer postoperative urinary tract infections in a propensity-matched analysis of a randomized trial.^22,23^ Both support the principle that topical delivery to an instrumented urinary compartment is feasible and effective. Whether a higher irrigant concentration would add further benefit is untested and merits prospective evaluation.

Systemic exposure appears to be minimal. Creatinine trajectories and acute kidney injury rates were indistinguishable between arms, and postoperative serum gentamicin was below the assay limit of quantitation in most sampled cases, with no evaluable sample above 0.6 µg/mL — under one-third of the trough threshold and one-twentieth of the peak threshold above which nephrotoxic and ototoxic risk rises.^24,25^ This is expected on mechanistic grounds. Urothelium is a barrier epithelium whose umbrella-cell tight junctions and apical uroplakin plaques exist to prevent solute movement between urine and blood, unlike the bowel and peritoneal surfaces across which the historical reports of aminoglycoside toxicity after surgical irrigation occurred.^26,27^ Consistent with this, no hypersensitivity, neuromuscular, or hearing-related events were observed, although audiometry was not performed and subclinical ototoxicity cannot be excluded.

From a stewardship standpoint, delivering the drug into the collecting system concentrates exposure where the organisms are rather than systemically. At full strength the irrigant sits well above the mutant selection window, the concentration range in which partially resistant subpopulations are preferentially enriched, so selection is unlikely while the irrigant runs.^28^

This study has several limitations. It was a retrospective single-center cohort study of nonrandomized, off-label treatment assignment at surgeon discretion, and residual confounding cannot be excluded. The arms were not fully concurrent, and although year of surgery, access modality, and surgeon were addressed analytically, unmeasured change in perioperative practice remains possible. The gentamicin arm contributed only three primary events, so subgroup and stratum-specific estimates are imprecise. A history of prior urinary infection was initially recorded incompletely and inconsistently and so could not contribute to the propensity score; however, any residual imbalance in that variable favors the control arm and was adjusted for in subsequent models. Upper-tract urine was not collected in standardized fashion before irrigation, precluding direct comparison of renal pelvic microbiology. Total irrigation volume was not recorded, so exposure could not be quantified per case. Finally, the economic analysis is modeled from published cost estimates rather than patient-level institutional accounting.

## Conclusion

In this matched cohort of 339 PCNL procedures, gentamicin-augmented intrarenal irrigation was associated with substantially lower 30-day urinary infectious morbidity, with no febrile urinary infection and no sepsis among treated patients. The association was robust to adjustment and across multiple sensitivity analyses. Short-term renal safety was comparable between groups. These findings support further prospective analysis of intrarenal gentamicin irrigation as a practical adjunct during PCNL.

## Data Availability

All data produced in the present work are contained in the manuscript

## Author Contributions

Study Concept and Design: Pengbo Jiang, Roshan M. Patel, Ryan S. Hsi

Data Acquisition: Kriselle Madamba, Jonathan Badin-Castro, Seyed Amiryaghoub Lavasani, Bruce M. Gao, Jocelyn Nguyen, Sohrab N. Ali, Pengbo Jiang

Analysis and Interpretation of Data: Pengbo Jiang, Jaylen M. Lee, Bruce M. Gao, Ryan S. Hsi Drafting of Manuscript: Pengbo Jiang, Ryan S. Hsi, Bruce M. Gao

Critical Revision: All authors

Supervision: Pengbo Jiang, Roshan M. Patel, Jaime Landman, Ralph V. Clayman

## Statements and Declarations

Ethical Approval and Consent to Participate: This study was conducted under an IRB-approved protocol at the University of California, Irvine (2016-2746). All procedures adhered to institutional guidelines and ethical standards consistent with the Declaration of Helsinki.

